# Brain-first forms of Parkinson’s Disease are over-represented in patients with non-responsive resting tremor

**DOI:** 10.1101/2024.07.23.24310859

**Authors:** Marcelo D. Mendonça, Pedro Ferreira, Raquel Barbosa, Joaquim Alves da Silva

## Abstract

Motor subtypes in Parkinson’s Disease (PD) are unstable over time, limiting mechanistic insights and biomarker discovery. We focused on Rest Tremor (RT) as a symptom to test for phenotype stability and link them to specific circuits and disease mechanisms. Using the PPMI cohort data over 5 years we found that RT we found that RT is more stable than common Tremor-Dominant definitions, a stability also seen for therapy response. At time of diagnosis, the population of therapy-resistant RT patients was enriched with a brain-first PD profile as predicted by a-Synuclein origin site and connectome (SOC) model. Resistant-RT patients have lower gastrointestinal and cardiovascular symptoms, lower prevalence of probable REM-Sleep behavior disorder, and higher dopaminergic asymmetry compared to therapy-responsive or no tremor patients. Treating RT as a distinct phenomenon revealed a relative phenotypic stability with treatment response being linked to different patterns of disease progression.

## Introduction

Motor subtypes in Parkinson’s Disease have been defined with algorithms using ratios between distinct motor symptom domains^1–4^. Individual subtypes (like Postural Instability/Gait Disorder, Akinetic-Rigid or Tremor-Dominant) have been described to be unstable^5,6^ over time limiting potential mechanistic understanding and biomarker discovery.

Across neuropsychiatric conditions, individual symptoms are expected to emerge from distinct brain circuits disorder^7^. Addressing symptoms as the relevant unit – avoiding classic conceptions that use severity ratios – may uncover more homogeneous clusters of PD patients. Therefore, symptom-based phenotyping can guide mechanistic studies, and link specific etiopathogenic mechanisms with topographic circuit involvement. A relative temporal stability could be an argument to support the causality of the mechanisms.

In this study we tested the temporal stability of PD phenotypes defined by Resting Tremor and its therapeutic response. Acknowledging the literature that relates therapy-resistant rest tremor with a benign PD prognosis^5,8^, we explored the additional hypothesis that patients presenting a therapy-resistant RT represent a population enriched in brain-first forms of PD. The a-Synuclein origin site and connectome (SOC) model proposes that anatomical location of the initial a-synuclein inclusion and propagation defines specific disease phenotypes^9^. This model has extensively been validated using non-motor aspects of the disease. Our study provides a proof-of-concept linking the presentation of a specific motor symptom with a framework of circuit involvement and disease mechanisms.

## Methods

### Subjects and Clinical Assessment

We accessed the Parkinson’s Progression Markers Initiative (PPMI) database (http://www.ppmi-info.org/) and extracted data regarding the first 5 years of follow-up of participants with a diagnosis of PD. OFF state evaluations were used for phenotypic stability assessment.

Motor ratings were obtained using the MDS-UPDRS. Part III of the MDS-UPDRS was used to score patients’ rigidity (total score on item 3.3), bradykinesia (total score on items 3.4, 3.5, 3.6, 3.7, 3.8, 3.9 and 3.14), action tremor (total score on items 3.15 and 3.16) and RT (total score on item 3.17). Patients were classified as RT present or absent based on the total score on item 3.17 (>0 or 0). Clinical status at year 2 was used as the reference year to study therapeutic response and for subgroup definitions. At same timepoint, total score of the part II of the MDS-UPDRS was used as well as the isolated point 2.10 (tremor). For Tremor-Dominant, we used previously defined criteria^10^

SCOPA-AUT (Scales for Outcomes in Parkinson’s Disease - Autonomic Dysfunction) subscores at baseline were used. Upper Gastrointestinal was defined by items 1-4; Lower Gastrointestinal by items 5-7; Urinary by 8-13; Cardiovascular by 14-16; Sexual by 22-25. Total score resulted from the sum of all points. MoCA (Montreal Cognitive Assessment) total score, as well as the % of patients with score < 26 were used.

Due to the non-linear relationship between tremor amplitude and likert-like scales, change in Tremor Amplitude were calculated using previously defined methods: %change = 100(10^0.5*(ScoreON – Score OFF)^-1).

## Data Analysis

For the Markov Chain Analysis 2 states were defined: Presence of RT or Absence of RT. Probability of transition was calculated yearly, and results were averaged. Labels were shuffled 1000 times to obtain a control distribution. Regarding stability of response, for each pair of consecutive years, the % of improvement was shuffled 1000 times between subjects to simulate a control distribution. Mean and a 95% Confidence Interval were calculated against which comparisons were performed. DaT imaging was obtained using single-photon emission computed tomography (SPECT) after ^123^I-FP-CIT intravenous injection. Binding ratios were extracted from those already processed centrally. Briefly, SPECT raw projection data was imported to a HERMES (Hermes Medical Solutions, Skeppsbron 44, 111 30 Stockholm, Sweden) system for reconstruction using the ordered subsets expectation maximization (OSEM) algorithm. This was done for all imaging centres to ensure the consistency of the reconstructions. Reconstruction was done without any filtering applied. The OSEM reconstructed files were then transferred to the PMOD (PMOD Technologies, Zurich, Switzerland) for subsequent processing. Attenuation correction ellipses were drawn on the images and a Chang attenuation correction was applied to images utilizing a site-specific μ that was empirically derived from phantom data acquired during site initiation for the trial. Once attenuation correction was completed a standard Gaussian 3D 6.0 mm filter was applied. These files were then normalized to standard Montreal Neurologic Institute (MNI) space so that all scans were in the same anatomical alignment. Next, the transaxial slice with the highest striatal uptake was identified and the 8 hottest striatal slices around it were averaged to generate a single-slice image. Regions of interest (ROI) were then placed on the left and right caudate, the left and right putamen, and the occipital cortex (reference region). Mean counts per voxel for each region were extracted and used to calculate binding potentials (BP) The absolute difference in binding or different regions (Full striatum, Putamen and Caudate) was calculated. Ratios to total binding were also calculated (Supplementary Figure 2). The distribution of asymmetries was performed for each analysis group and the difference of these distributions with the reference distribution (Resistant Tremor) was calculated.

### Magnetic Resonance Imaging dataset

The PPMI’s magnetic resonance imaging (MRI) dataset was used to extract, when available, patients’ T1-weighted images. These images were generated with a 1.5–3 Tesla scanner and acquired as a 3D sequence in the three main axis (axial, sagittal and coronal) ^11^. Only baseline images were selected. A total of 373 images were extracted.

### MRI-based extraction of amygdala volumes

Amygdala volumes were extracted from T1-weighted images using the preprocessing pipeline of voxel-based morphometry (VBM), available through Computer Anatomy Toolbox (CAT12, v12.8.2), running on Statistical Parametric Mapping (SPM12, v7771)^12^. Standard VBM preprocessing protocols and default parameters of CAT12 were used. Images were axially pre-aligned along the anterior/posterior commissure plane. Briefly, T1-weighted images were denoised and bias-corrected for intensity inhomogeneities, and then segmented into grey matter (GM), white matter (WM) and cerebrospinal fluid (CSF) following SPM’s unified segmentation ^13^ through tissue probability maps. The segmented tissue maps were then spatially normalized to the standard Montreal Neurological Institute (MNI-152) space with a voxel size of 1.5 × 1.5 × 1.5 mm^3^, and an atlas-based extraction of region of interest (ROI) volumes (i.e. amygdala volumes) was conducted automatically through regional labelling based on the Neuromorphometrics atlas implementation in CAT12 ^12^ (figure 2E). To account for possible brain atrophy or lesions, ROI-based volumes were normalized to an estimation of brain parenchyma volume, computed as the sum of total grey and white matter volumes.

Descriptive statistics were presented as means and standard deviations. When two groups with continuous variables were compared either Mann-Whitney test was used. Spearman’s rank correlation coefficient was used. For 3 groups, Kruskall-Wallis test with Dunn post-hoc test was used. For OFF/ON comparisons, Mixed-Effect models with post-hoc Sidak multiple comparison test was used. For categorical variables, the Chi-square test was used. The significance level was set at 0.05.

## Results

### Phenotypes based on Rest Tremor presence and therapeutic response are relatively stable and may define distinct subgroups

Using data from the first 5-year follow-up of the PPMI cohort we found that the presence or absence of RT (defined by the OFF assessment) was relatively stable across time (Fig.1A-B). A Markov chain analysis of transition state (939 unique subjects at baseline) revealed that the probability of yearly stability in category of patients with RT was significantly higher than what would be expected by chance (88% vs 67.4% ± 1.7), a similar result was seen for patients with no RT (73% vs 32.8% ± 2.5). Stability was lower when classical TD definitions^10^ were used (∼78%, Fig.1B) conducting to the classical decrease in TD phenotypes (Fig.1C) vs. PIGD increase across time (Supplementary Figure 1A).

**Figure 1.**
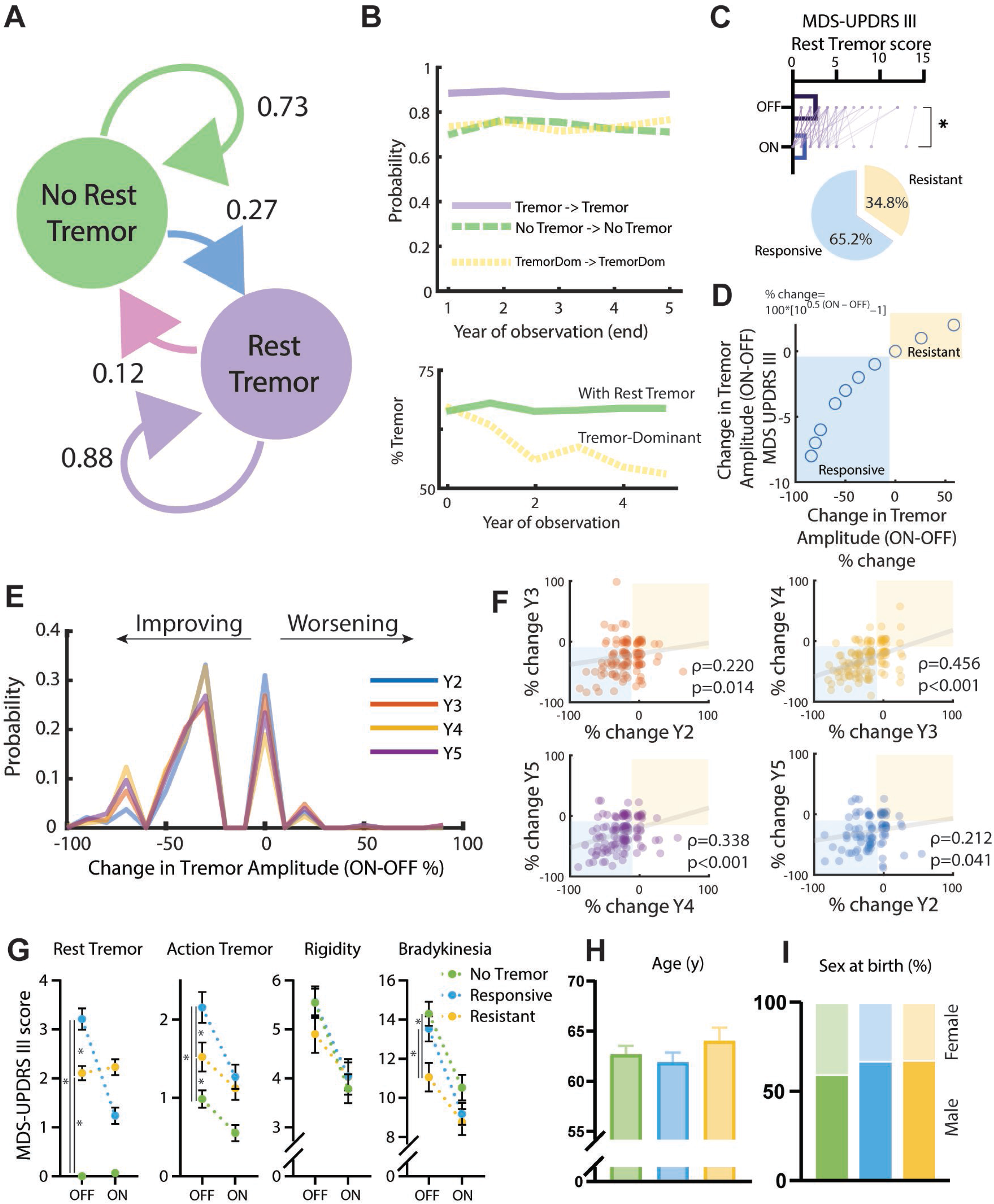
Rest Tremor phenotypes relatively stable during the first 5 years of disease. A) Classifying PD patients with patients with Rest Tremor (RT) and with No Rest Tremor across the 5 years of disease progression revealed that patients with RT have a high probability of maintaining RT across this period. B) Top: Year-to-year probability of maintaining the same-classification. Tremor-dominant definition used previously defined algorithms. Bottom: Fraction of patients presenting RT at the off state across 6 observation points and fraction of patients classified as Tremor-Dominant (Baseline – Year 0 – to Year 5). C) RT scores in OFF state are higher than in the ON state at Y2 (the first point with this data available), even facing high variability. Bottom: 34.8% of patients do not present a therapeutic response in RT. D) Responses in tremor amplitude do not behave linearly. E) Probability distribution on change in RT responses in different time points present a similar pattern. F) Correlation between tremor responses in consecutive years, and between Y2 and Y5. G) Clinical assessment extracted from MDS-UPDRS III at Y2 of 4 symptoms (rest tremor, action tremor, rigidity and bradykinesia) in the OFF and ON assessment dividing across phenotypes (No Tremor, Tremor-resistant, Tremor responsive). Results from Mixed Effect Models. Rest Tremor: ON/OFF F(1,289)=99.0, p<0.001; Group: F(2,328)=104.7, p<0.001; ON/OFFxGroup: F(2,289)=155.3; Post-hoc: No Tremor vs. Responsive: p<0.001; No Tremor vs. Resistant: p<0.001; Responsive vs Resistant: p<0.001. Action Tremor: F(1,289)=53.52, p<0.001; Group: F(2,328)=12.93, p<0.001; ON/OFFxGroup: F(2,289)=7.46, p<0.001; Post-hoc: No Tremor vs. Responsive: p<0.001; No Tremor vs. Resistant: p=0.0346; Responsive vs Resistant: p=0.0021. Rigidity: ON/OFF F(1,286)=130.7, p<0.001; Group: F(2,325)=0.2363, p=0.7897; ON/OFFxGroup: F(2,286)=3.658, p=0.027; Post-hoc No Tremor vs. Responsive: p=0.9836; No Tremor vs. Resistant: p=0.1915; Responsive vs Resistant: p=0.1979. Bradykinesia: ON/OFF F(1,288)=219.4, p<0.001; Group: F(2,327)=3.356, p=0.0361; ON/OFFxGroup: F(2,288)=5.975, p=0.0029; Post-hoc No Tremor vs. Responsive: p=0.3452; No Tremor vs. Resistant: p=0.0011; Responsive vs Resistant: p=0.0150. H) Age at assessment, One Way ANOVA: F(2,328)=0.890, p=0.4117. I) Sex at birth: Chi-Square=2.08, p=0.35

**Figure 2.**
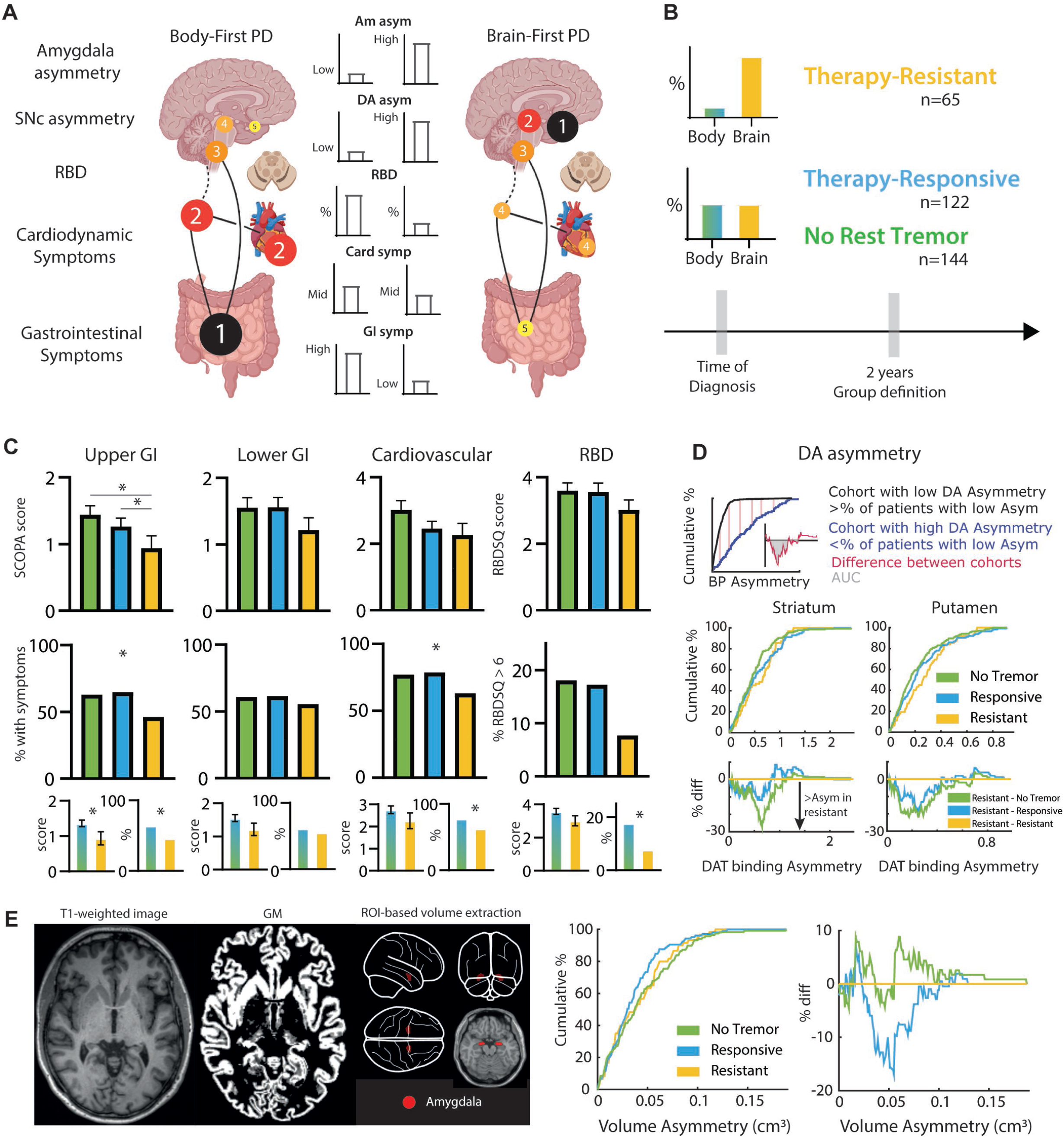
The population of treatment Resistant RT is enriched with brain-first forms of PD. A) The a-Synuclein origin site and connectome (SOC) model predicts 2 main pathways of disease progression: Brain First and Body First forms. We expect Body First patients to present at time of motor symptoms onset (4 in the left part), involvement of Gastrointestinal, Cardiac and brainstem areas related to REM-Sleep Behaviour disorder. By comparison, at time of symptoms onset (2 in the right part) we expect brain first patients to not present pathological involvement of these systems. B) At time of diagnosis (motor symptoms onset) we expect that groups enriched with brain-first phenotypes (including therapy-resistant RT) present a distinct profile of symptoms vs. the body first phenotypes. C) Top: SCOPA score and RBDSQ scores for each symptom domain. Upper Gastrointestinal – Kruskall Wallis F(2,330)=7.708, p=0.0212, post-hoc: No tremor vs. Improvement: p=0.6251; No tremor vs. resistant p= 0.0066; Improvement vs. persistant: p=0.0243; Lower Gastrointestinal – Kruskall Wallis F(2,330)=1.941, p=0.3788; Cardiovascular – Kruskall Wallis F(2,330)=3.623; RBDSQ – Kruskall Wallis F(2,331)=1.290, p=0.5246; Middle: % of patients with SCOPA subdomains score > 1 and RBDSQ-score > 6; Upper Gastrointestinal – Chi-square = 6.860, p=0.0324; Lower Gastrointestinal – Chi-square = 0.7295 p=0.6944; Cardiovascular – Chi-square = 6.067 p=0.0481; RBD – Chi-square = 3.962, p=0.1379. Bottom: SCOPA and RBDSQ scores and paired % of patients with SCOPA subdomains score > 1 and RBDSQ > 6. Upper Gastrointestinal – Mann-Whitney, p=0.0061; Chi-square = 6.305, p=0.0120; Lower Gastrointestinal – Mann-Whitney, p=0.1816; Chi-square = 0.718, p=0.3967; Cardiovascular – Mann-Whitney, p=0.0752; Chi-square = 5.958, p=0.0147; RBD – Mann-Whitney, p=0.2791; Chi-square = 3.962, p=0.0475; D) Asymmetry in binding potential of DaT-Scan. Top: Example of the used metrics of dopaminergic asymmetry based on the cumulative distribution in the cohort. Black line: Example distribution from a cohort with low asymmetry; Blue line: Example distribution from a cohort with high asymmetry. Red line, difference between cohorts. Middle: Cumulative distribution of Striatal and Putaminal binding potential differences in the 3 cohorts. Bottom: Difference in distribution asymmetry between pairs of subgroups taking the resistant group as a reference. AUC for Striatal Binding Potential in Tremor-resistant > Tremor-responsive: 57.69%; > No Tremor: 85.66%; AUC for Putamen Binding Potential in Tremor-resistant > Tremor-responsive: 81.46%; > No Tremor: 95.96%; E) Asymmetry in Amygdala volumes. Left: Example of a T1-weighted image from a PPMI subject and corresponding grey matter segmentation, spatially normalized to the MNI-152 common template. Representation of atlas-based regions of interest for left and right amygdala, overlayed on a schematic brain and a normalized T1-weighted image. Middle: Cumulative distribution of amygdala volumes differences in the 3 cohorts. Right: Difference in distribution asymmetry between pairs of subgroups taking the resistant group as a reference. AUC for amygdala asymmetry Tremor-resistant > Tremor-responsive: 95.2%; > No Tremor: 38.9%; * p<0.05.

After 2 years of follow-up, clinical data from 424 subjects with PD was available. Hundred and forty-four (34%) subjects had no RT while 280 (66%) presented RT when assessed in the OFF state. Assessment on a ON state was available for 187 of patients with RT (67%) and a significant decrease in RT on the ON state was observed (Fig.1C) even if individual variability in responses was clear. At Y2 approximately 1/3 of patients with RT did not improve their tremor score on the ON state (Fig.1C). The relationship between tremor amplitude and clinical scores is non-linear manner, so we used previously established methods to quantify response ^14^ (Fig.1D). Importantly, even with loss to follow-up, the distributions of individual improvement in tremor response were comparable across 4 timepoints (Y2, Y3, Y4 and Y5, Fig.1E) and when the OFF-ON response difference was compared across time (i.e., across contiguous years of assessment), an association between therapeutic responses magnitude were found (Fig.1F). Response to treatment across years was also more similar than what would be randomly expected (Supplementary Figure 1B) suggesting a relative stability of RT therapy-response phenotype.

Patients were then divided into 3 groups based on the Y2 phenotype: No Rest Tremor, Therapy-responsive tremor and Therapy-resistant tremor. No significant differences were seen in gender (Fig.1I) and age (Fig.1H, although therapy-resistant tremor patients tended to be older). Therapy had an effect in action tremor, bradykinesia and rigidity reduction across all groups (Fig.1G). Importantly, the therapy-resistant group had a significantly lower OFF state bradykinesia score.

### Patients with therapy-resistant rest tremor have an increase in features suggestive of Brain-first forms of PD

The SOC model proposes that for the same “severity” of involvement of the dopaminergic system, brain-first cases are expected to present lower gastrointestinal, cardiac and rates of REM-Sleep behavior disorders (RBD) than body-first cases. The model also predicts higher asymmetry on dopaminergic and amygdala integrity (Figs.2AB).

Using the data at time of diagnosis, we found that patients with therapy-resistant RT presented a lower rate and severity of upper GI symptoms as measured by the SCOPA, a lower rate of cardiovascular symptoms and a trend for a lower number of positive RBD screening cases (Fig.2C) when compared with the other 2 groups. When power was increased by joining the 2 comparison groups, the previous comparisons remained significant, with a trend in lower GI symptoms and a significant lower number of RBD screening cases in therapy-resistant group (Fig.2C). Importantly, lower scores in the therapy-resistant group were not found in MDS-UPDRS II assessment (Supplementary Figure 2A), urinary, sexual or total SCOPA score (Supplementary Figure 2B) or MoCA score (Supplementary Figure 2C), suggesting symptom-specificity, more than a general reduction in self-reported symptoms or other dimensions.

Asymmetries in Putaminal Binding Potential (as assessed by quantitative DaT-SPECT) revealed that the therapy-resistant group had a higher frequency of more asymmetric patients than therapy-responsive or no tremor patients (Fig.2D). These results were seen whether the full striatum or only the putamen were compared, and across normalization methods (Supplementary Figure 2D). Results on amygdala volumes were less clear – although tremor-resistance was associated to higher asymmetry than tremor responsiveness, this was not seen for the no-tremor group (Fig.2E). These results not seen in other brain structures (Supplementary Figure 2E) and were maintained after normalization to brain volume (Supplementary Figure 2F).

## Discussion

Contrary to what is seen with classical phenotyping, treating patients with RT as an individual group allow us to document a relative phenotypic stability in the first 5 years of disease. RT is a distinct symptom in PD, that contrary to bradykinesia, is not present in all patients. Although DA depletion is necessary for RT to emerge, heterogeneity of RT responses to DA replacement exist and its association with depletion severity remains unclear^15^. The emergence of RT must involve changes in specific topographies of the nervous system that lead to the of the 4-6 Hz oscillations. In the neurodegenerative context this must involve damage to specific circuits. Reconsidering available data on prion-like propagation of synuclein^16^ it is tempting to link disease progression based on structural connectivity, brain circuits involvement and development of specific symptoms. If the disease process starts in distinct regions in different subjects, connectivity-based progression would lead to a distinct set of symptoms.

Classical phenotyping and natural history assessment has revealed that Tremor-Dominant forms of PD have a slower progression, and better prognosis^8^. There is an additionally entity – Benign tremulous parkinsonism (BTP)^17–20^ – with neuropathologically confirmed Lewy Bodies and nigral cell loss^19^ who have long and benign courses and whose tremor has minimal levodopa responsiveness (i.e, they are therapy-resistant). The SOC model makes specific predictions that can causally link tremor and benign progression.

We found that patients with a therapy-resistant RT have clinical features suggesting an enrichment with brain-first disease forms. This group had a lower rate of RBD, upper GI and cardiovascular symptoms and higher rates of asymmetric dopaminergic degeneration. This associates therapy-resistant rest tremor with the dysfunction pattern specific of brain-first disease^9^. A last observation is that tremor-resistant patients present a significantly lower bradykinesia score and a trend for an older age of disease onset. This argues in favor of a particular “more benign” phenotype with characteristics akin to the ones described in BTP.

Our analysis has limitations. Data were extracted from a naturalistic study, where formal levodopa challenge tests with supramaximal dosages were not performed. Therapeutic responses include multiple agents (Levodopa, DA agonists and/or anti-cholinergic) and a bias related to more aggressive treatment of more severe forms could not be excluded. Nevertheless, this analysis is in line with previous results suggesting that in 1/3 of PD patients, RT does not respond to supramaximal LD replacement therapy^21^. Importantly, potential variability that could lead to a non-differential misclassification of clinical subpopulations is expected to bias results towards the null. Our positive results emerge even with this limitation. Although non-motor symptoms were assessed using self-reported scales, significant differences were only found in *a priori* defined domains and not in global SCOPA, urinary or sexual scores suggesting domain specificity. This is reinforced by the lack of differences in global cognition, global MDS-UPDRS part II (motor experiences of daily living) but a distinct response in self-reported tremor.

Although conflicting evidence exists, the pattern of dopamine loss in tremor^19–21^ and therapy-resistant tremor^19,22–24^PD phenotypes seem to be distinct from other forms. This SNc topographical sparing could be caused by two, non-mutually exclusive mechanisms: 1) Individual cell-specific resilience to death^25^ (i.e., polymorphisms or mutations that reduce vulnerability to specific cell populations) and 2) connectivity related patterns of cell damage^26^. Our data suggest that this second mechanism may have a role on motor phenotypes. This can happen by a distinct pattern of SNc degeneration but also by differential extra-nigral circuits pathology.

## Data Availability

The PPMI data is part of an open database. Access to PPMI data can be requested at
https://www.ppmi-info.org/access-data-specimens/download-data.

## Acknowledgements

Data used in the preparation of this article were obtained on May, 13, 2023 from the Parkinson’s Progression Markers Initiative (PPMI) database (https://www.ppmi-info.org/access-data-specimens/download-data), RRID:SCR_006431. This analysis used data openly available from PPMI. For up-to-date information on the study, visit http://www.ppmi-info.org. PPMI – a public-private partnership – is funded by the Michael J. Fox Foundation for Parkinson’s Research and funding partners, including 4D Pharma, Abbvie, AcureX, Allergan, Amathus Therapeutics, Aligning Science Across Parkinson’s, AskBio, Avid Radiopharmaceuticals, BIAL, BioArctic, Biogen, Biohaven, BioLegend, BlueRock Therapeutics, Bristol-Myers Squibb, Calico Labs, Capsida Biotherapeutics, Celgene, Cerevel Therapeutics, Coave Therapeutics, DaCapo Brainscience, Denali, Edmond J. Safra Foundation, Eli Lilly, Gain Therapeutics, GE HealthCare, Genentech, GSK, Golub Capital, Handl Therapeutics, Insitro, Jazz Pharmaceuticals, Johnson & Johnson Innovative Medicine, Lundbeck, Merck, Meso Scale Discovery, Mission Therapeutics, Neurocrine Biosciences, Neuron23, Neuropore, Pfizer, Piramal, Prevail Therapeutics, Roche, Sanofi, Servier, Sun Pharma Advanced Research Company, Takeda, Teva, UCB, Vanqua Bio, Verily, Voyager Therapeutics, the Weston Family Foundation and Yumanity Therapeutics. We thank João Duarte, PhD for the feedback in volumetric analysis.

## Data availability

The PPMI data is part of an open database. Access to PPMI data can be requested at https://www.ppmi-info.org/access-data-specimens/download-data.

## Author contributions

MM designed the study, conducted all analysis and redacted the first draft of the manuscript with feedback from JAS. PF performed the MRI analysis. RB critically reviewed the manuscript and analysis. JAS provided significant input on data analysis, interpretation and manuscript redaction. All authors contributed to the final manuscript redaction and read and approved the final manuscript.

