## Supplemental for "Brain-first forms of Parkinson’s Disease are over-represented in patients with non-responsive resting tremor"

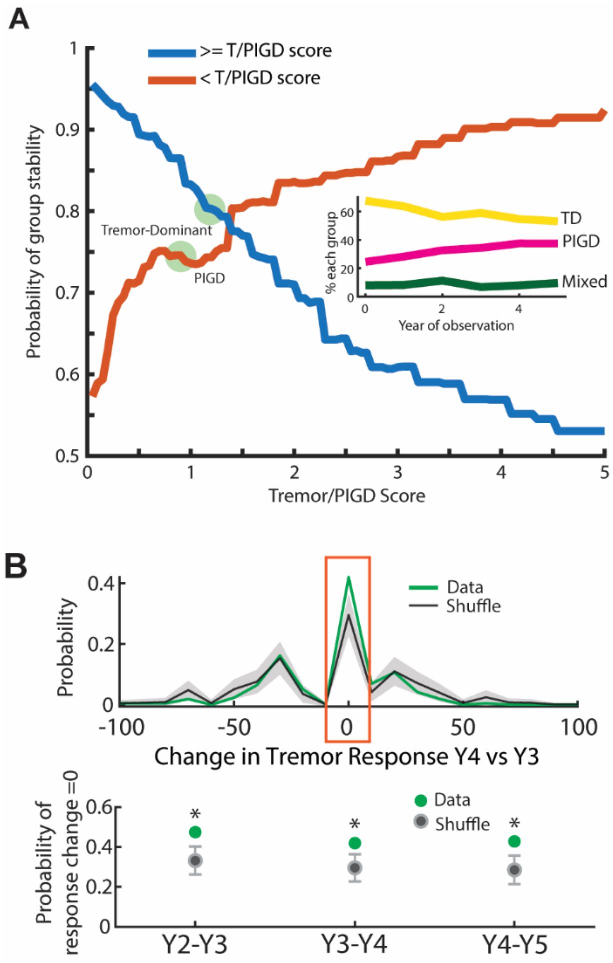


**Supplementary Figure 1 –** A) Simulation of stability results for different thresholds of Tremor/PIGD score in the definition of Tremor-Dominant & PIGD scores. Circles: Criteria typically used. Changing thresholds will change stability, but with significant changes in the number of patients included in each group. Inset: Percentage of subjects classified in each category. A decrease in TD patients is accompanied by a progressive increase in the PIGD classification. B) Top: Distribution of the differences in the individual probability of responses between Y4 and Y3 and shuffle control. Bottom: Probability of no change in therapeutic response is significantly higher that randomly expected across timepoints. Black line represents the average of simulated sham distributions and shaded area represents the 95% Confidence Interval of this distribution. * $p<0.05$ vs. shuffle

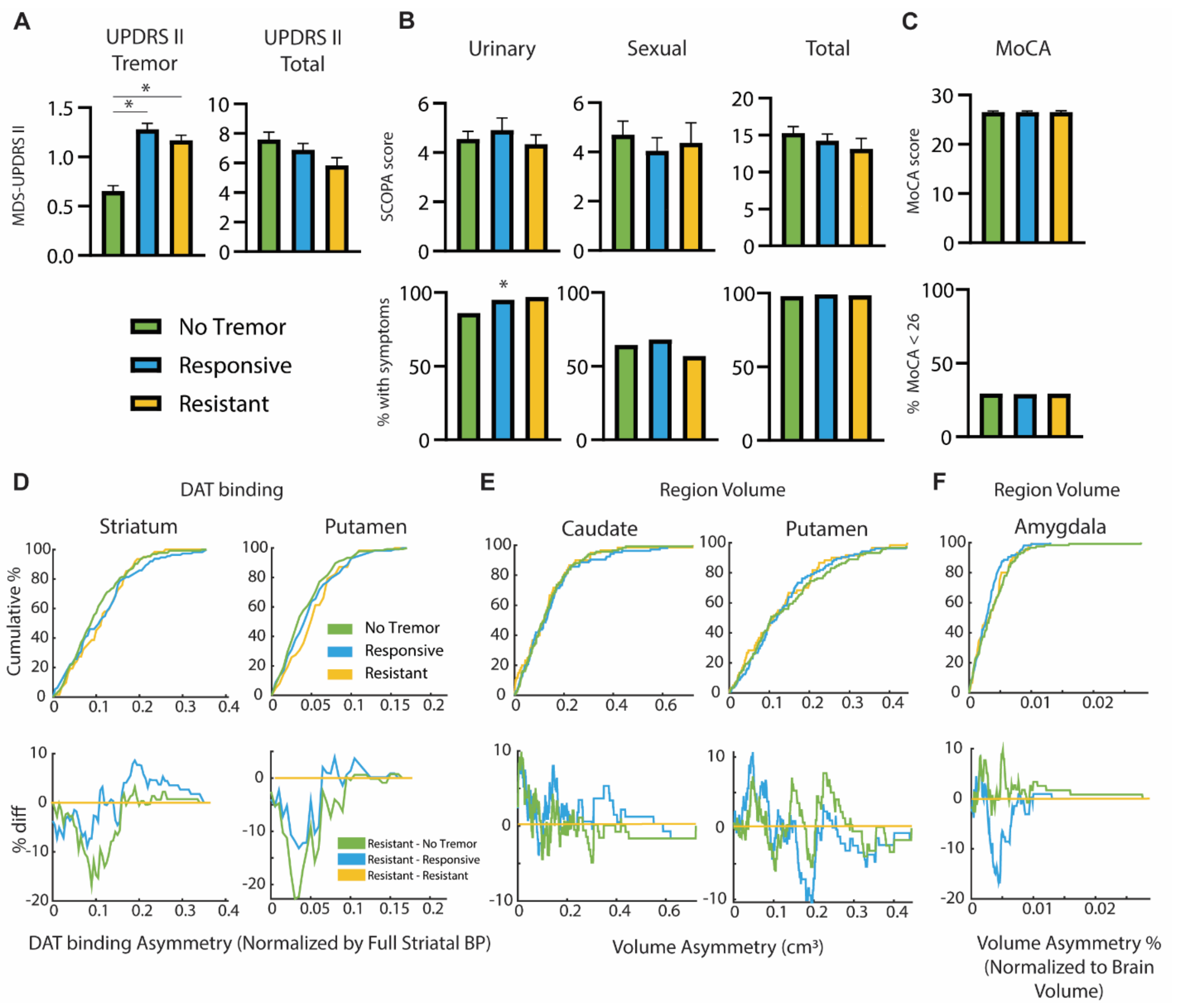


**Supplementary Figure 2 –** A) MDS-UPDRS Part II Scores; Tremor – Kruskall Wallis, F(2,331)=64.58, p<0.001, post-hoc: No tremor vs. Improvement: p<0.001; No tremor vs. resistant p<0.001; Improvement vs. persistent: p=0.5604; MDS-UPDRS Part II Total: – Kruskall Wallis, F(2,330)=3.483, p=0.1752; B) Top: SCOPA score for each symptom domain. Urinary – Kruskall Wallis F(2,330)=0.1773, p=0.9164; Sexual – Kruskall Wallis F(2,330)=0.5651, p=0.7539; Total – Kruskall Wallis F(2,330)=3.865, p=0.1448; Bottom: % of patients with SCOPA subdomains score > 1 Urinary – Fisher Exact Test, p=0.0083; Sexual – Chi-Squared=2.279, p=0.3199; Total – Fisher Exact Test p>0.999; C) Top: MOCA score - Kruskall Wallis F(2,329)=0.7783, p=0.6776, Bottom: % of patients with MOCA < 26: Chi-Squared=0.0156, p=0.9922; D) Cumulative distribution of Striatal and Putaminal binding potential, normalized to total BP in the 3 cohorts. E) Cumulative distribution of caudate and putamen volumes asymmetry. F) Cumulative distribution of amygdala volume asymmetry, normalized to total volume of gray and white matter in the 3 cohorts.
